# Coping under stress: Prefrontal control predicts stress burden during the COVID-19 crisis

**DOI:** 10.1101/2021.06.28.21259570

**Authors:** Maximilian Monninger, Tania M. Pollok, Pascal-M. Aggensteiner, Anna Kaiser, Iris Reinhard, Andrea Hermann, Andreas Meyer-Lindenberg, Daniel Brandeis, Tobias Banaschewski, Nathalie E. Holz

**Author notes:** Corresponding author, **Address for correspondence:** Dr. Nathalie E. Holz, Department of Child and Adolescent Psychiatry and Psychotherapy, Central Institute of Mental Health, Medical Faculty Mannheim / Heidelberg University, J 5, 68159 Mannheim, Germany, Tel: +49 621/1703-4904, Fax: +49 621/1703-1205.

## Abstract

**Background:** The coronavirus (COVID-19) pandemic has confronted millions of people around the world with an unprecedented stressor, affecting physical and mental health. Accumulating evidence suggests that emotional and cognitive self-regulation is particularly needed to effectively cope with stress. Therefore, we investigated the predictive value of affective and inhibitory prefrontal control for stress burden during the COVID-19 crisis.

**Method:** Physical and mental health burden were assessed using an online survey, which was administered to 104 participants of an ongoing German at-risk birth cohort during the first wave in April 2020. Two follow-ups were carried out during the pandemic, one capturing the relaxation during summer and the other the beginning of the second wave of the crisis. Prefrontal activity during emotion regulation and inhibitory control were assessed prior to the COVID-19 crisis.

**Results:** Increased inferior frontal gyrus activity during emotion regulation predicted lower stress burden at the beginning of the first and the second wave of the crisis. In contrast, inferior and medial frontal gyrus activity during inhibitory control predicted effective coping only during the summer, when infection rates decreased but stress burden remained unchanged. These findings remained significant when controlling for sociodemographic and clinical confounders such as stressful life events prior to the crisis or current psychopathology.

**Conclusions:** We demonstrate that differential stress-buffering effects are predicted by the neural underpinnings of emotion regulation and cognitive regulation at different stages during the pandemic. These findings may inform future prevention strategies to foster stress coping in unforeseen situations.

**Highlights:**

- Health threatening stressors, such as the COVID-19 pandemic, significantly worsen well-being.
- Results reveal high levels of stress during the course of the pandemic with an increase of stress burden towards the second wave.
- Self-regulation is an important coping strategy to restore allostasis.
- Higher prefrontal activity during emotion regulation predicted less stress during the peaks of infection rates in the first and second wave
- Higher prefrontal inhibitory control predicted less stress burden between both waves when infection rates were low.
- Our findings highlight the importance of prefrontal regulation as effective coping mechanisms in the face of unprecedented stressors.

## 1. Introduction

The coronavirus (COVID-19) pandemic is related to physical health impairments and dramatic changes to everyday life for hundreds of millions of people around the world. With the social contact restrictions beginning in March 2020 in Germany, nearly all schools were closed overnight, working environments changed radically, and social life was restricted probably as never before. Early studies investigating the impact of these initial “first-level threats” (imminent threat) on mental health burden reported elevated levels of perceived stress, anxiety, and depressive symptoms during the first wave of the COVID-19 pandemic (Benke, Autenrieth, Asselmann, & Pané-Farré, 2020a, 2020b). While sustained socio-economic “second-level threats” continued throughout the time when infection rates lowered during the summer, renewed first-level health threats emerged at the beginning of the second wave of the pandemic in late 2020.

Appropriate coping in response to these time-dependent challenges in the face of an ongoing crisis like the COVID-19 pandemic requires the balanced use of emotional regulation and cognitive control strategies (Feder, Fred-Torres, Southwick, & Charney, 2019). Indeed, so far, several studies investigated the effect of emotion regulation strategies and cognitive control on well-being and mental health during the pandemic. Findings indicate that adaptive emotion regulation strategies predicted increased well-being, whereas maladaptive emotion regulation strategies and deficits in cognitive control predicted higher levels of anxiety, greater depressive symptoms, reduced quality of life, risky health behavior, and diminished well-being during the pandemic (Appelhans, Thomas, Roisman, Booth-LaForce, & Bleil, 2021; Breaux et al., 2021; Brehl, Schene, Kohn, & Fernández, 2021; Low, Overall, Chang, & Henderson, 2020; Panayiotou, Panteli, & Leonidou, 2021; Weissman et al., 2021; Yang, Liu, Li, & Shu, 2020). While affective control, such as reappraisal, involves the adaptive modulation of emotions in response to unpleasant stimuli (Gross, 2015; Holley, Ewing, Stiver, & Bloch, 2017), cognitive control includes the inhibition of inappropriate or ineffective behavior (Aron, 2007). Both are part of the executive function system (Holley et al., 2017) and reflected mainly in the prefrontal cortex (PFC; Davidson, Putnam, & Larson, 2000; Ochsner, Bunge, Gross, & Gabrieli, 2002). Affective control strategies are associated with an increased activity in the inferior frontal gyrus (IFG), the ventrolateral PFC, dorsolateral PFC, and medial PFC (Gross, 2015; Kanske, Heissler, Schonfelder, Bongers, & Wessa, 2011; Ochsner et al., 2002). Cognitive inhibitory control is linked to activity in the fronto-basal-ganglia circuit including the middle frontal gyrus (MFG), the IFG, and the basal ganglia (Aron, 2007; Aron, Fletcher, Bullmore, Sahakian, & Robbins, 2003; Aron, Robbins, & Poldrack, 2004; Verbruggen & Logan, 2008), indicating shared neuronal underpinnings within the IFG and MFG.

The present longitudinal study investigated the predictive value of prefrontal activity during an emotion regulation and an inhibitory control task assessed prior to the COVID-19 pandemic for stress burden during the COVID-19 crisis in 104 participants of an ongoing at-risk cohort study following participants since birth. Approximately four weeks after the initial lockdown in Germany in March 2020, participants rated their current stress burden in an online survey (baseline during the first wave). Six to eight weeks later (first follow-up) as well as six months later (at the beginning of the second wave of the pandemic; second follow-up), participants completed the same questionnaire. We expected that higher affective and cognitive control predict lower stress burden during the COVID-19 pandemic. Given the ongoing uncertainties about the course and duration of the pandemic and its unprecedented nature, we had no specific a-priori hypotheses about how neural activity predict stress burden at different stages of the pandemic. However, based on previous findings (Brehl et al., 2021; Weissman et al., 2021; Yang et al., 2020), we speculate that the ability to control emotions in order to overcome perceived stress burden would be highly necessary at the beginning of the first wave of the COVID-19 crisis, when participants were confronted with direct threat due to high infection and death numbers (first-level threat). By contrast and based on previous findings assessed during the course of the pandemic (Appelhans et al., 2021), we anticipated that cognitive control would become increasingly important in order to cope with the ongoing uncertainties during the pandemic (second-level threat).

## 2. Materials and methods

### 2.1 Sample

The present investigation was conducted within the Mannheim Study of Children at Risk, an ongoing longitudinal study of the long-term outcomes of early psychosocial and biological risk factors following participants since birth (Laucht et al., 2000). The initial sample consisted of 384 children born between 1986 and 1988 in the Rhine-Neckar region of Germany. Infants were recruited from two obstetric and six children’s hospitals and were included according to a two-factorial design intended to enrich and control the risk status of the sample (distribution in the current sample: 37 (35.6%) participants without psychosocial risk, 35 (33.7%) with low psychosocial risk, and 32 (30.8%) with high psychosocial risk at birth).

Up to the time of the lockdown on March 23^rd^ 2020, 165 participants completed an ongoing assessment wave, including a comprehensive questionnaire package on physical and mental health, a structured clinical interview (SCID-5-CV German version; (Beesdo-Baum, Zaudig, & Wittchen, 2019)), and a magnetic resonance imaging (MRI) session, including an emotion regulation and stop-signal task (see below). In April 2020 (during the first wave), two months later (first follow up during summer), and six months later (second follow-up in December), participants were invited to take part in an online COVID-19 questionnaire, adapted and modified from the Coronavirus Health and Impact Survey (CRISIS, (Nikolaidis et al., 2020)). 133 (80.61%) participants completed the COVID-19 baseline assessment, of whom 128 (96.24%) also responded to the first follow-up questionnaire and 116 (87.21%) to the second follow-up questionnaire. Functional MRI data were available for 104 participants for the baseline and first follow-up questionnaire, and for 95 participants for the second follow-up questionnaire. The study was approved by the Ethics Committee of the University Heidelberg, Germany, written informed consent from all participants was obtained, and participants were financially compensated.

### 2.2 Stress burden during COVID-19

To assess the impact and threat of COVID-19 on physical and mental health, we used four items (‘The impact of COVID-19 on my physical health is?’, ‘COVID-19 is a potential threat to my physical health’) and mental health (‘The impact of COVID-19 on my mental health is?’, ‘COVID-19 is a potential threat to my mental health’) rated on a 10-point Likert scale (0: completely disagree; 10: completely agree). We calculated a sum score for each assessment (baseline, first follow-up, second follow-up).

### 2.3 Emotion Regulation

#### 2.3.1 Emotion regulation Task

Prior to the COVID-19 pandemic, participants performed an adapted and modified version of an emotion regulation task (Hermann, Kress, & Stark, 2017) during functional MRI. In brief, participants were asked to either watch aversive (‘Look_negative_’) or neutral (‘Look_neutral_’) pictures from the International Affective Picture System (IAPS; (Lang, Bradley, & Cuthbert, 1999)) or to reappraise negative pictures (‘Reappraisal’). In the reappraisal condition, participants were asked to use the strategy of reappraisal to decrease the intensity of their negative affect. The participants were carefully instructed to either view the depicted scenario from a more positive or at least a less negative point of view (e.g. a person in jail might be a famous actor) or to rationalize the presented picture (e.g. due to enormous advances in modern medicine, a very premature baby may have an entirely normal life). During the neutral conditions, participants were instructed to simply watch the depicted scenarios without actively changing their emotional state evoked by the pictures.

The task consisted of a randomized block design, in which every block started with a jittered 3s presentation of the instruction form (i.e. ‘Look’ or ‘Reappraise’). Subsequently, participants viewed either four negative or four neutral pictures for 5s each according to the presented condition, and were asked to rate the intensity of currently perceived negative feelings on a 7-point Likert scale (1 = no negative feelings at all; 7 = extremely negative feelings) via a button press (max 4s). Subsequently, a white fixation cross on black background was presented during the inter-trial-interval up to a total block duration of 30s. The total task comprised four blocks per condition (12 blocks in total) and lasted for 6 min 37s. The blocks were arranged in four runs with a randomized presentation of all conditions within each run, leading to a maximum of two presentations of the same condition in succession.

### 2.4 Inhibitory control

#### 2.4.1 Inhibitory control Task

Participants completed the stop-signal task (SST) to assess cognitive inhibitory control during functional MRI (Rubia, Smith, Brammer, & Taylor, 2003). Each trial began with a fixation cross (500ms), which was followed by an arrow pointing to the left or right (go-signal). In the go-trials (75% of a total number of 160 trials), participants were instructed to react to the arrow as quickly and accurately as possible by pressing either the left or the right button according to the previously shown arrow. Stop-trials (25% of a total number of 160 trials) were designed as trials in which a go-signal was followed by an arrow pointing upwards (stop-signal). During the stop trials, participants were instructed to inhibit their response (yielding successful vs. unsuccessful inhibition). The delay between a go-signal and a stop-signal (stop-signal delay; SSD) started at 250ms and was dynamically changed according to the participant’s performance using an adaptive algorithm. The SSD latency increased by 50ms whenever the participant correctly inhibited the previous response (max. 900ms), making it more difficult to stop. In contrast, the SSD latency decreased by 50ms whenever the previous stop-trial was incorrectly answered (min. 50ms). Using this procedure, an approximately equal number of successful and unsuccessful stop-trials can be achieved (inhibitory control in the present sample was on average 60.79 % (SD = 12.68)). Moreover, we excluded participants with a negative stop-signal reaction time (SSRT). Prior to scanning, all participants were given clear instructions. Total scanning time was 6 min 37s.

### 2.5 Functional MRI data acquisition and preprocessing

Functional MRI data collection consisted of a localizer scan followed by a blood oxygen level-dependent (BOLD)-sensitive T2*-weighted echo-planar imaging (EPI) sequence and structural T1-weighted sequence using a 3T-scanner (PrismaFit; Siemens) with a standard 32-channel head coil. For functional imaging, a total of 186 volumes with 36 slices covering the whole brain (matrix 64 x 64, resolution 3.0 x 3.0 x 3.0 mm with 1 mm gap, repetition time = 2100 ms, echo time = 35 ms, flip angle = 90°) were acquired for each task. The slices were inclined 20° from the anterior/posterior commissure level. The first 11 volumes of the emotion regulation task and the first six volumes of the SST were discarded to allow longitudinal magnetization to reach equilibrium.

### 2.6 Functional MRI data analyses and statistical analyses

Statistical parametric mapping version 12 (SPM12) implemented in MATLAB R2017b was used to analyze functional MRI data. Preprocessing included slice time correction of the volumes to the first slice, realignment to correct for movement artifacts, co-registration of functional and anatomical data, spatial normalization to standard Montreal Neurological Institute (MNI) space, and spatial smoothing with a Gaussian filter of 8 mm full-width at half maximum. Motion parameters were examined to ensure head movement did not exceed 3 mm.

For the emotion regulation task, individual first-level contrasts based on onsets and durations of each condition were convolved with the canonical hemodynamic response function (HRF) in order to model the BOLD time course by using general linear models. Six motion parameters were included as regressors of no interest. For the group-level analysis, individual contrast images of the Reappraisal > Look_neutral_ condition were entered into a random-effects analysis to assess emotion regulation.

Similarly, for the SST, individual first-level contrasts for the onsets of the four event types (correct go-trials, incorrect go-trials, successful inhibition, and unsuccessful inhibition) were modeled using delta functions convolved with the canonical HRF. Again, six motion estimation parameters were included as regressors of no interest. Second-level random effects analyses were modeled for the contrast of interest, i.e. successful inhibition was determined by contrasting successful stop-trials with unsuccessful inhibition and correct go-trials [successful inhibition > (unsuccessful inhibition – correct go-trials)]. Whole-brain differences in activation for the contrasts of interest for both tasks were estimated using t-tests at a family-wise error (FWE) corrected p-value of .05.

The IFG and the MFG were defined as regions of interest (ROIs) based on previous studies reporting a strong involvement in emotion regulation and inhibitory control, and their potential relevance in coping under stress (Berretz, Packheiser, Kumsta, Wolf, & Ocklenburg, 2021; Gavazzi, Giovannelli, Curro, Mascalchi, & Viggiano, 2020; Golde et al., 2020; Rosenbaum et al., 2018). To extract predefined ROIs, anatomical masks implemented in the Wake Forest University (WFU) PickAtlas v2.4(Maldjian, Laurienti, Kraft, & Burdette, 2003) were used, where a p < 0.05 FWE correction (minimum of 10 adjacent voxels) was applied. Mean contrast values of each participant were extracted from the FWE-corrected significant clusters in the contrasts of interest (Table 1) and exported to SPSS Statistics 25. The predictive power of IFG and MFG activity during affective and inhibitory control for the impact of COVID-19 on mental and physical health was investigated using linear regression models controlling for all covariates (see below). Moreover, a conservative Bonferroni correction was applied for the hemispheres, two regions of interest (MFG and IFG), and for affective and cognitive control (α of 0.05 divided by 8 tests, resulting in α_adj_=.0063). Additional sensitivity analyses were performed to control for the robustness of significant effects.

**Table 1:**
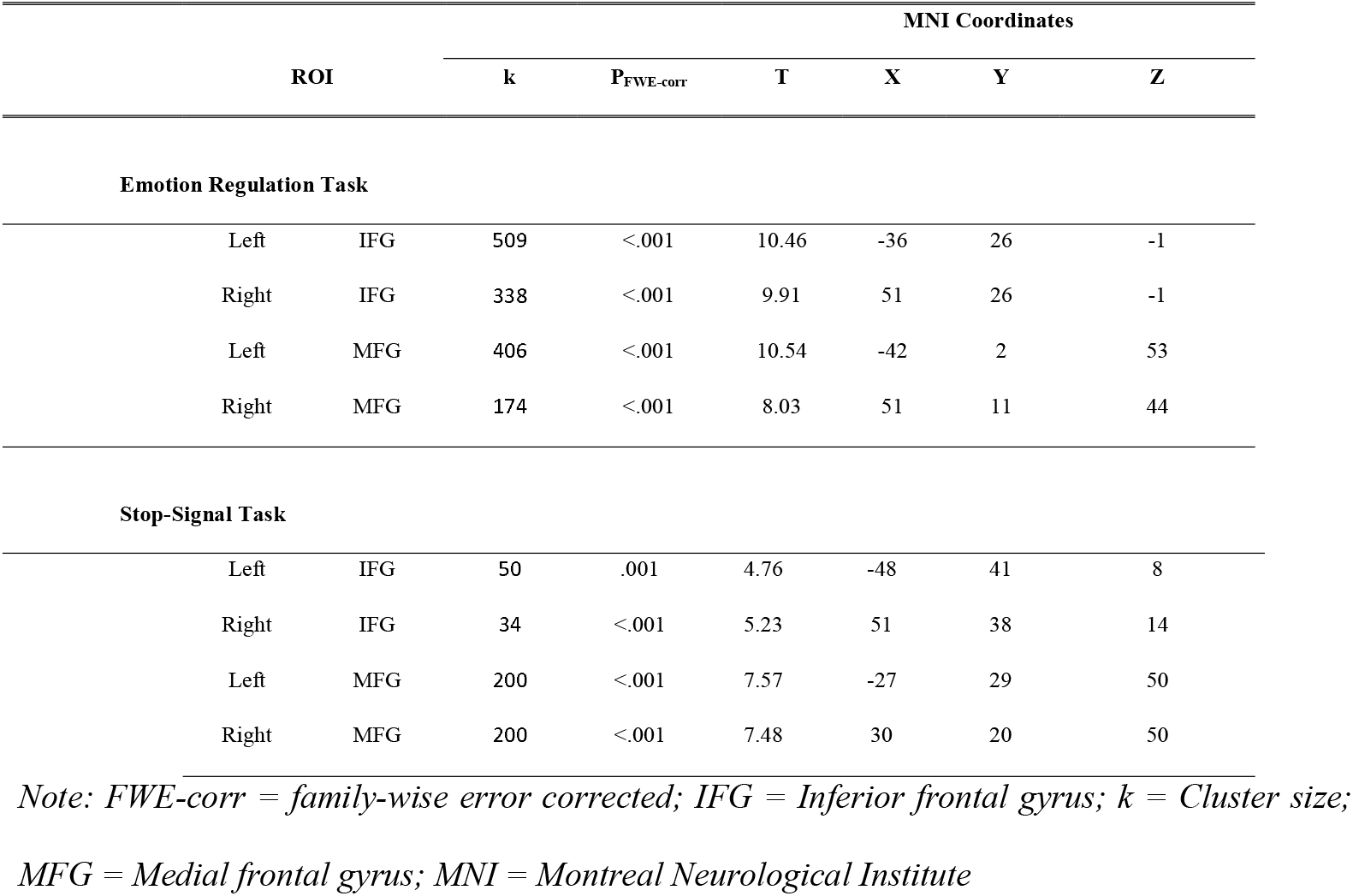
Results of the ROI analyses for the emotion regulation task and the stop-signal task

### 2.7 Covariates

Several important covariates were added to the main analyses, including gender, psychosocial adversity and obstetric risk at birth, current diagnoses of a mental disorder (assessed prior to the pandemic), as well as current life events prior to and during the COVID-19 pandemic.

#### 2.7.1 Psychosocial adversity

Psychosocial adversity was assessed using a standardized parent interview according to an enriched family adversity index (Rutter & Quinton, 1977) at the participants’ age of 3 months. The interview comprised 11 items covering characteristics of the family environment, the parents, and the parents’ partnership (e.g., presence of parental psychiatric disorders, overcrowding in the home, ongoing parental conflicts, or unwanted pregnancy) during a period of one year prior to the assessment. A sum score of psychosocial adversities was calculated by adding up the presence of all items.

#### 2.7.2 Obstetric risk at birth

Obstetric adversity was assessed using a standardized parent interview conducted at the participants’ age of 3 months. A sum score of obstetric risk factors was computed by adding up the presence of nine adverse conditions during pregnancy, delivery, and in the early postnatal phase, such as preterm birth or low birth weight (Laucht et al., 2000).

#### 2.7.3 Life events

Life events were recorded using a modified version of the Munich Events List (MEL) (Maier-Diewald, 1983) within the online questionnaire at all three time points. The MEL covers several areas of acute and chronic, positive and negative stressors, including marriage, delivery of a child, but also negative health outcomes, illness of a relative or job loss. We adjusted the items depending on the time point, i.e., presence of life events between the regular assessment wave and prior to the COVID-19 crisis for the COVID-19 baseline assessment, and life events between baseline assessment and first follow-up as well as between the first and second follow-up for both COVID-19 follow-up assessments, respectively.

### 2.8 Sensitivity analyses

Further sensitivity analyses were calculated to control for the robustness of the predictive value of the neural activity, including parenthood, household income, current work status, workplace changes, and whether the participants were critical workers for the COVID-19 response as additional covariates.

## 3. Results

### 3.1 Descriptive data and changes of COVID-19 impact

Descriptive data are depicted in Table 2. There were significant differences for the reported physical and mental impact of the COVID-19 pandemic between the baseline assessment and the follow-up assessments (F = 11.812, P < .001). Post-hoc analyses revealed no significant differences between the baseline assessment and the first follow-up (P = .894), but significant differences emerged between the baseline and the second follow-up (T = - 4.306, P < .001), and between the first and second follow-up (T = -3.912, P <.001), suggesting a continuous burden on mental and physical health since the beginning of the COVID-19 outbreak, with a further increased stress burden at the beginning of the second wave of the pandemic.

**Table 2:**
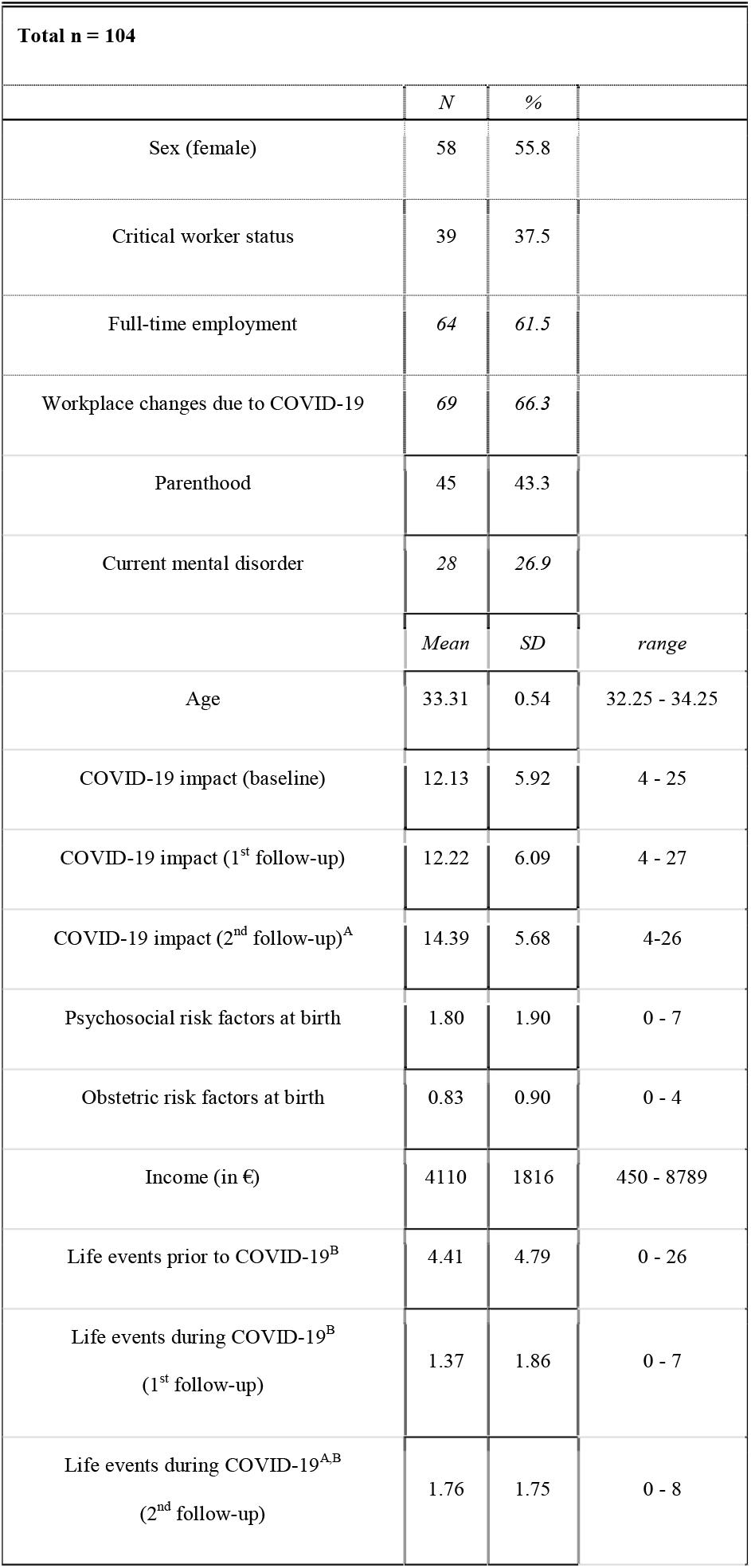
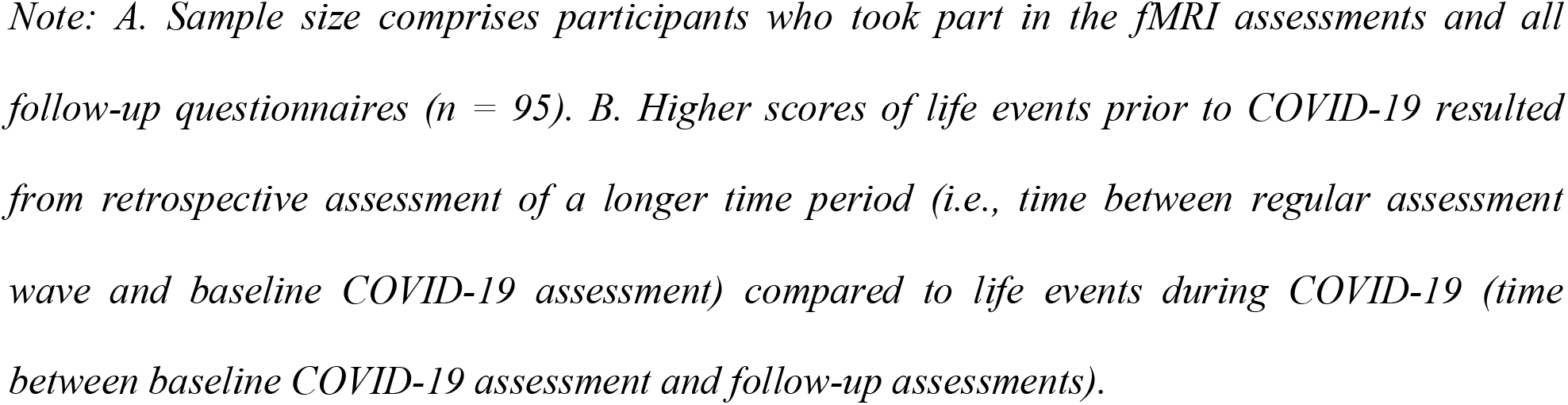
Sample description and descriptive data

| Total n = 104 |  |  |  |
| --- | --- | --- | --- |
|  | *N* | *%* |  |
| Sex (female) | 58 | 55.8 |  |
| Critical worker status | 39 | 37.5 |  |
| Full-time employment | 64 | 61.5 |  |
| Workplace changes due to COVID-19 | 69 | 66.3 |  |
| Parenthood | 45 | 43.3 |  |
| Current mental disorder | 28 | 26.9 |  |
|  | *Mean* | *SD* | *range* |
| Age | 33.31 | 0.54 | 32.25 - 34.25 |
| COVID-19 impact (baseline) | 12.13 | 5.92 | 4 - 25 |
| COVID-19 impact (1<sup>st</sup> follow-up) | 12.22 | 6.09 | 4 - 27 |
| COVID-19 impact (2<sup>nd</sup> follow-up)<sup>A</sup> | 14.39 | 5.68 | 4-26 |
| Psychosocial risk factors at birth | 1.80 | 1.90 | 0 - 7 |
| Obstetric risk factors at birth | 0.83 | 0.90 | 0 - 4 |
| Income (in €) | 4110 | 1816 | 450 - 8789 |
| Life events prior to COVID-19<sup>B</sup> | 4.41 | 4.79 | 0 - 26 |
| Life events during COVID-19<sup>B</sup><br>(1<sup>st</sup> follow-up) | 1.37 | 1.86 | 0 - 7 |
| Life events during COVID-19<sup>A,B</sup><br>(2<sup>nd</sup> follow-up) | 1.76 | 1.75 | 0 - 8 |

*Note: A. Sample size comprises participants who took part in the fMRI assessments and all follow-up questionnaires (n = 95). B. Higher scores of life events prior to COVID-19 resulted from retrospective assessment of a longer time period (i.e., time between regular assessment wave and baseline COVID-19 assessment) compared to life events during COVID-19 (time between baseline COVID-19 assessment and follow-up assessments).*

### 3.2 COVID-19-related stress burden at the baseline assessment

Higher right IFG activity during the emotion regulation task was associated with a lower stress burden at the baseline assessment (β = -.281, T = -3.023, P = .003, Figure 1a), after controlling for gender and psychosocial and obstetric risk factors at birth, current mental disorder prior to the COVID-19 pandemic, and current life events. In contrast, left IFG and bilateral MFG activity during emotion regulation were not related to stress burden at the beginning of the COVID-19 crisis (P >.121). In addition, inhibition-related right MFG activity was negatively associated with the COVID-19 burden at the baseline assessment (β = -.215, T = -2.220, P = .029), although this was not significant after correction for multiple testing. No significant relationships emerged with regard to left MFG and IFG activity during inhibition.

**Figure 1:**
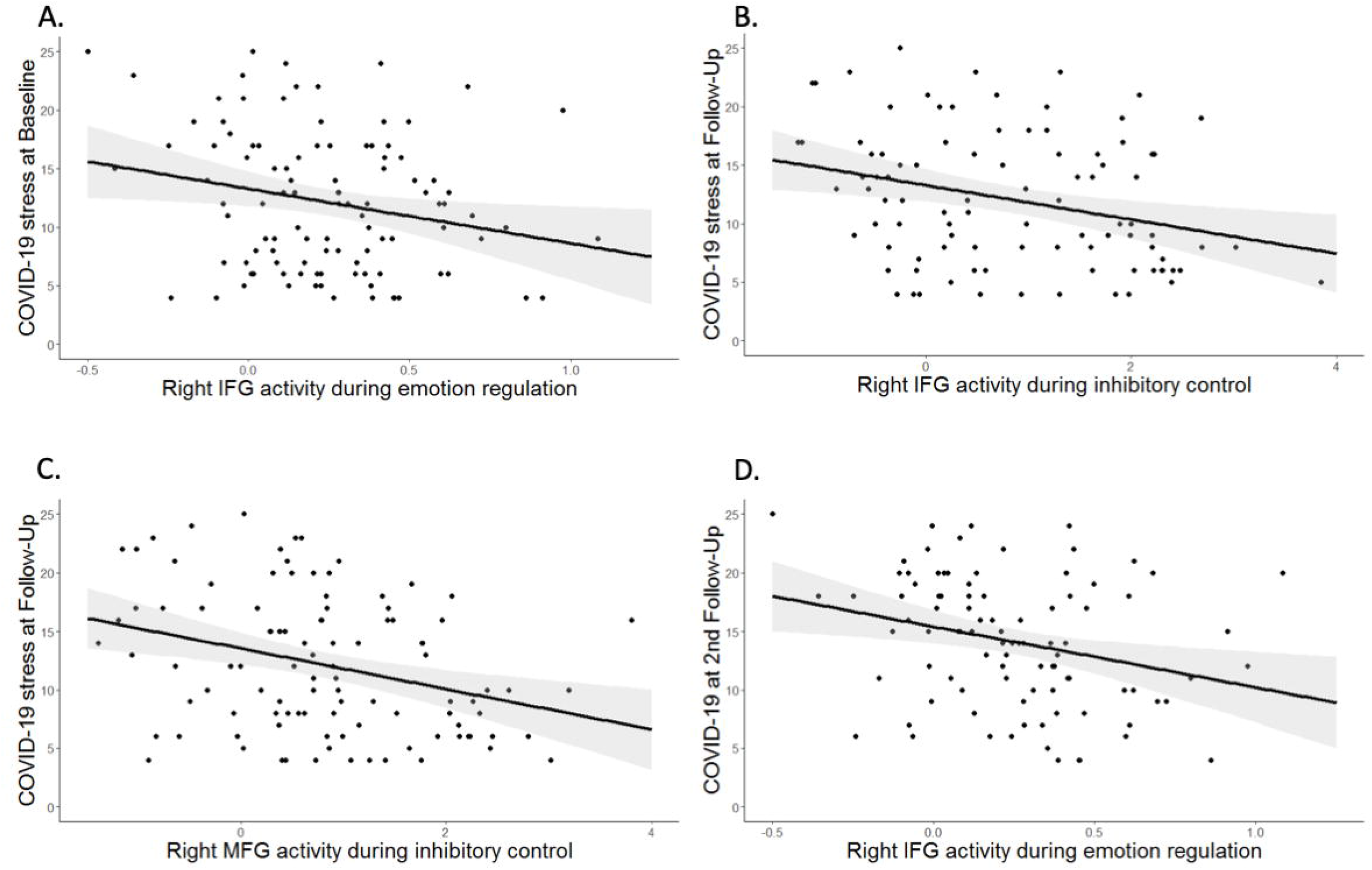
A. Association of right IFG activity and COVID-19 impairments at the baseline assessment. Higher affective control was related to decreased stress burden during the first wave of the COVID-19 pandemic. B. and C. Association of right IFG activity and right MFG activity and COVID-19 impairments at the first follow-up assessment during the summer. Higher cognitive activity was related to decreased stress burden caused by the COVID-19 pandemic. D. Association of right IFG activity and COVID-19 impairments at the second follow-up assessment. Higher affective control was related to decreased stress burden at the beginning of the second wave of the pandemic.

### 3.3 COVID-19-related stress burden at the first follow-up assessment

During the first follow-up assessment in the summer, neither IFG nor MFG during emotion regulation was related to the COVID-19 impact (all Ps > 0.61), suggesting that affective control did not predict stress burden at this point of time. In contrast, lower right IFG and right MFG activity during inhibitory control was associated with a higher COVID-19 stress burden at the first follow-up assessment, even when controlling for the above-mentioned covariates (right IFG: β =-.348, T=-3.638, P<.001; right MFG: β =-.375, T=-3.884, P<.001, Figure 1b&1c). Moreover, a negative association for left MFG activity emerged, which did not survive correction for multiple comparisons after including the above-mentioned covariates (β =-.250, T=-2.582, P=.011). Inhibition-related left IFG activity was not related to stress burden at the first follow-up (P = .093).

### 3.4 COVID-19-related stress burden at the second follow-up assessment

Higher right IFG activity during the emotion regulation task was associated with a lower stress burden at the second follow-up assessment (β = -.327, T = -3.246, P = .002, Figure 1d), after controlling for the above-mentioned covariates. In contrast, left IFG and MFG activity during emotion regulation were not related to stress burden during the second wave of the COVID-19 pandemic (all Ps >.086). Likewise, inhibition-related activity was not related to COVID-19 burden at the second follow-up (all Ps > .105).

### 3.5 Sensitivity analyses

Separate subsequent sensitivity analyses were calculated including additional control variables such as parenthood, income, current work status, critical worker status, and workplace changes due to the COVID-19 pandemic. None of the sensitivity analyses changed the results (Table 3).

**Table 3:** Results of further sensitivity analyses using brain activity as main predictor to explain stress burden.

| covariate | ROI | Task / Assessment | Beta | T | p-value |
| --- | --- | --- | --- | --- | --- |
| **Parenthood** | Right IFG | Emotion Regulation / Baseline | -.259 | -2.681 | **.009** |
|  | Right MFG | Stop-Signal Task / Follow-Up | -.333 | -3.428 | **.001** |
|  | Right IFG | Stop-Signal Task / Follow-Up | -.313 | -3.288 | **.001** |
|  | Right IFG | Emotion Regulation / 2<sup>nd</sup> Follow-Up | -.325 | -3.203 | **.002** |
| **Income** | Right IFG | Emotion Regulation / Baseline | -.288 | -2.997 | **.003** |
|  | Right MFG | Stop-Signal Task / Follow-Up | -.351 | -3.574 | **.001** |
|  | Right IFG | Stop-Signal Task / Follow-Up | -.294 | -2.973 | **.004** |
|  | Right IFG | Emotion Regulation / 2<sup>nd</sup> Follow-Up | -.339 | -3.404 | **.001** |
| **Current work status** | Right IFG | Emotion Regulation / Baseline | -.275 | -2.853 | **.005** |
|  | Right MFG | Stop-Signal Task / Follow-Up | -.375 | -3.866 | **< .001** |
|  | Right IFG | Stop-Signal Task / Follow-Up | -.319 | -3.268 | **.001** |
|  | Right IFG | Emotion Regulation / 2<sup>nd</sup> Follow-Up | -.311 | -3.182 | **.002** |
| **Critical worker status** | Right IFG | Emotion Regulation / Baseline | -.271 | -2.858 | **.005** |
|  | Right MFG | Stop-Signal Task / Follow-Up | -.367 | -3.765 | **< .001** |
|  | Right IFG | Stop-Signal Task / Follow-Up | -.306 | -3.113 | **.002** |
|  | Right IFG | Emotion Regulation / 2<sup>nd</sup> Follow-Up | -.324 | -3.221 | **.002** |
| **Workplace changes** | Right IFG | Emotion Regulation / Baseline | -.271 | 2.803 | **.006** |
|  | Right MFG | Stop-Signal Task / Follow-Up | -.363 | -3.702 | **< .001** |
|  | Right IFG | Stop-Signal Task / Follow-Up | -.311 | -3.148 | **.002** |
|  | Right IFG | Emotion Regulation / 2<sup>nd</sup> Follow-Up | -.323 | -3.225 | **.002** |

## 4. Discussion

To the best of our knowledge, this is the first study to report a differential predictive value of emotion- and inhibition-related prefrontal control for stress burden at the beginning of and during the COVID-19 crisis, respectively (Figure 2).

**Figure 2:**
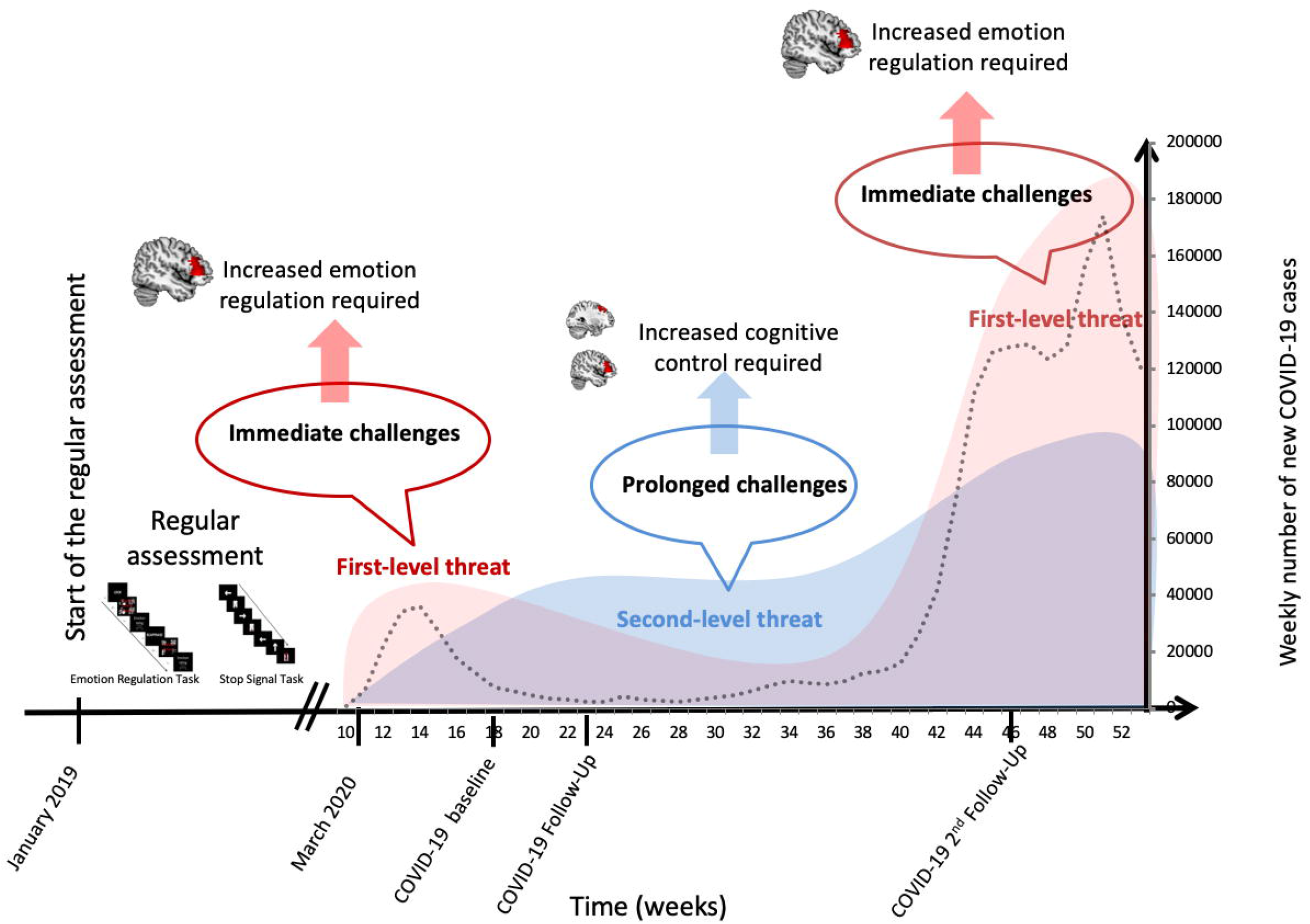
Timeline and proposed stress model at different stages during the COVID-19 pandemic. An initial high load of emotional distress requires adequate affective coping of the IFG at the beginning of the first and the second wave of the COVID-19 pandemic. Prolonged socio-economic uncertainties involve cognitive control of the IFG and MFG to overcome these challenges during the ongoing crisis. *Note:* Weekly statistics on COVID-19 cases are freely available from the Robert Koch Institute, Berlin, Germany.

Specifically, we found a significant negative relationship between right IFG activity during emotion regulation assessed prior to the COVID-19 pandemic and stress burden approximately four weeks after the lockdown in Germany. In light of unforeseen first-level threat entailing immediate danger to health and emotional challenges due to the dramatic, unprecedented social contact restrictions at the beginning of the crisis with high levels of emotional arousal, fear and the imminent feeling of loneliness (Bauerle et al., 2020; Brooks et al., 2020; Dubey et al., 2020; Odriozola-Gonzalez, Planchuelo-Gomez, Irurtia, & de Luis-Garcia, 2020), affective coping was particularly needed (Groarke et al., 2020; Restubog, Ocampo, & Wang, 2020; Weissman et al., 2021). Our results support this assumption, indicating that participants who are characterized by higher affective control were able to cope better with these initial threats and uncertainties of an unprecedented stressor like the COVID-19 pandemic. At the first follow-up assessment, which occurred during the summer, when the first-level threat decreased due to loosened restrictions and lower infection rates. Also, emotional habituation may occur if participants are confronted with the same stressor for a longer amount of time (Grissom & Bhatnagar, 2009). These reasons both may contribute to a decreased necessity for affective coping. Notably, towards the second wave of the pandemic, when infection rates increased dramatically even above those during the first wave and a second lockdown was imposed, affective coping was again required to face this immediate, even more intense, threat.

In contrast, we found an increased need for cognitive control strategies when facing second-level threat, i.e. socio-economic uncertainties. Specifically, higher cognitive inhibition-related activity in the right MFG and IFG predicted lower COVID-19 distress only at the first follow-up (when restrictions had eased), but was unrelated to stress burden at the beginning of the first wave (baseline) and the second wave (second follow-up). While there was an immediate threat at the beginning of both the first and the second wave, the progression of the pandemic is characterized by ongoing socio-economic challenges, uncertainties about one’s own future financial and work capabilities, reduced working hours, and the length of the crisis (Cutler & Summers, 2020; Fegert, Vitiello, Plener, & Clemens, 2020). These sustained and secondary challenges (Pedrosa et al., 2020) require increased cognitive coping and flexibility over time, thus explaining the negative association of the MFG and IFG activity with stress burden at the first follow-up.

Given the unique prospective design of our study, we were able to demonstrate the strong predictive value of neural activity for stress coping during the pandemic, irrespective of the presence of several important factors which have previously been reported to affect stress coping. Specifically, the findings were robust against control for the presence of other current stressful life events (Undheim & Sund, 2017), socio-economic disadvantages (McEwen & Gianaros, 2010), the occurrence of early life psychosocial adversities (Quinlan et al., 2017; Sheffler, Piazza, Quinn, Sachs-Ericsson, & Stanley, 2019), or the presence of a mental disorder before the crisis (Joormann & Gotlib, 2010), thus highlighting the superior role of neural self-regulation for coping under stress.

Our results provide a substantial contribution to the existing literature, and have important implications not only for affective and cognitive coping with potential further waves of increasing COVID-19 cases, for instance due to virus mutations around the world, but also with unprecedented stressful events in general. Specifically, we argue that there is a particular need for prevention strategies which aim at improving the individual’s affective and cognitive coping capacities in response to unforeseen events with a high stress load. As such, we suggest that stepped care neuromodulation might mitigate stress burden via gaining self-control over neural affective and cognitive regulation. Therefore, enhancing neuroplasticity through different methods, such as mindfulness or even neurofeedback in emotion regulation areas might offer the potential to acquire and foster primary preventive self-regulation skills when initially encountering stressful events, whereas learning self-control over cognitive regulation activity might rather serve as a secondary prevention tool during ongoing stressful events. In addition, ecological momentary assessment (EMA) might be a promising, affordable tool to further foster affective and cognitive control strategies in response to real-time, real-life stressors on an individual’s own smartphone; this could be addressed by future intervention studies.

Some limitations of our findings need to be addressed. Only a third of our initial sample was able to take part in all COVID-19 assessments. Moreover, despite the quick start, we were unable to include the first days of the lockdown, which might have exerted the greatest effects on perceived stress. However, since we were particularly interested in coping with stress burden caused by the crisis, this time period appears to be appropriate to capture stress burden given that previous studies found no differences when comparing well-being, anxiety and depression in the initial phase of the pandemic to four weeks later (Vindegaard & Benros, 2020).

In the framework of an ongoing longitudinal study following at-risk participants since birth, we thus provide first evidence for the predictive value of neural underpinnings of emotion regulation during the first and second lockdown and cognitive regulation during the COVID-19 pandemic between both waves. These findings may inform future prevention strategies seeking to foster stress coping in unforeseen situations.

## Data Availability

Data is available upon reasonable request.

## Financial support

NH and TB gratefully acknowledge grant support from the German Research Foundation (grant numbers DFG HO 5674/2-1, GRK2350/1) and the Ministry of Science, Research and the Arts of the State of Baden-Württemberg, Germany (Sonderfördermaßnahme SARS CoV-2 pandemic to NH). The funding source had no role in study design, collection, analysis, and interpretation of data, in the writing of the report, and in the decision to submit the article for publication.

## Conflicts of Interest

T.B. served in an advisory or consultancy role for ADHS digital, Infectopharm, Lundbeck, Medice, Neurim Pharmaceuticals, Oberberg GmbH, Roche, and Takeda. He received conference support or speaker’s fee by Medice and Takeda. He received royalities from Hogrefe, Kohlhammer, CIP Medien, Oxford University Press; the present work is unrelated to these relationships. D.B. serves as an unpaid scientific consultant for an EU-funded neurofeedback trial unrelated to the present work. A.M.-L. has received consultant fees from the American Association for the Advancement of Science, Atheneum Partners, Blueprint Partnership, Boehringer Ingelheim, Daimler und Benz Stiftung, Elsevier, F. Hoffmann-La Roche, ICARE Schizophrenia, K. G. Jebsen Foundation, L.E.K Consulting, Lundbeck International Foundation (LINF), R. Adamczak, Roche Pharma, Science Foundation, Sumitomo Dainippon Pharma, Synapsis Foundation – Alzheimer Research Switzerland, System Analytics, and has received lectures fees including travel fees from Boehringer Ingelheim, Fama Public Relations, Institut d’investigacions Biomèdiques August Pi i Sunyer (IDIBAPS), Janssen-Cilag, Klinikum Christophsbad, Göppingen, Lilly Deutschland, Luzerner Psychiatrie, LVR Klinikum Düsseldorf, LWL Psychiatrie Verbund Westfalen-Lippe, Otsuka Pharmaceuticals, Reunions i Ciencia S. L., Spanish Society of Psychiatry, Südwestrundfunk Fernsehen, Stern TV, and Vitos Klinikum Kurhessen.

## Author Contributions

M.M. conducted the MRI data collection, analyzed all data and wrote the paper. T.M.P. collected and analyzed the data and reviewed the paper. P.-M.A. collected the data and reviewed the paper. A.K. reviewed the paper. I.R. supervised all data analyses and reviewed the paper. A.H. contributed to the design of the study and reviewed the paper. A.M.-L. and D.B. contributed to the design of the study, supervised data analyses and reviewed the paper. T.B. obtained funding for the study, supervised data analyses and reviewed the paper. N.E.H. obtained funding for the study, designed the study, supervised all data analyses and wrote the paper.

## Acknowledgements

The authors gratefully thank Manfred Laucht (1946-2020), who was one of the founders of the Mannheim Study of Children at Risk and who continuously acted as an inspiring and supporting mentor giving impulses for innovative research projects. M.M. and N.H. had full access to all of the data in the study and take responsibility for the integrity of the data and the accuracy of the data analysis. The authors thank Sibylle Heinzel, Rafaela Gehr, Christin Loebel and Cäcilia Pracht for conducting and supporting the assessments.

## Notes

### Author Declarations

The study was approved by the Ethics Committee of the University Heidelberg, Germany, written informed consent from all participants was obtained, and participants were financially compensated.

